# An extension of the Benefit Risk Assessment of VaccinEs (BRAVE) Toolkit to evaluate Comirnaty and Spikevax vaccination in the European Union

**DOI:** 10.1101/2024.08.09.24311669

**Authors:** Neilshan Loedy, Hector G Dorta, Steven Abrams, Jonas Crèvecoeur, Daniel R. Morales, Catherine Cohet, Lander Willem, Geert Molenberghs, Niel Hens, Xavier Kurz, Chantal Quinten, Johan Verbeeck

## Abstract

**Introduction:** Amid the global COVID-19 pandemic, vaccines were conditionally authorised for human use to protect against severe infection. The BRAVE toolkit, a user-friendly R Shiny application, was developed retrospectively together with the European Medicine Agency (EMA) with the aim of fulfilling the need for flexible tools to assess vaccine benefits and risks during and outside a pandemic situation.

**Method:** This study employed BRAVE to evaluate the impact of COVID-19 mRNA vaccines across 30 EU/EEA countries by quantifying the number of prevented clinical events (i.e., confirmed infections, hospitalisations, intensive care unit (ICU) admissions, and deaths), using a probabilistic model informed by real-time incidence data and vaccine effectiveness estimates. The analysis assumes fixed population dynamics and behaviour. Additionally, BRAVE assesses risks associated with mRNA-based vaccines (myocarditis or pericarditis) by comparing observed incidence rates in vaccinated individuals with background incidence rates.

**Results:** mRNA vaccines were estimated to directly prevent 11.150 million (95% Confidence Interval (CI): 10.876 – 11.345) confirmed COVID-19 infections, 0.739 million (95% CI: 0.727 – 0.744) COVID-19 hospitalisations, 0.107 million (95% CI: 0.104 – 0.109) ICU admissions, and 0.187 million (95% CI: 0.182 – 0.189) COVID-19-related deaths in the EU/EEA between 13 December 2020 and 31 December 2021. Despite increased vaccination-associated myocarditis or pericarditis observed in younger men, the benefits of vaccination still outweigh these risks.

**Conclusion:** Our study supports the benefit/risk profile of COVID-19 vaccines and emphasises the utility of employing a flexible toolkit to assess risks and benefits of vaccination. This user-friendly and adaptable toolkit can serve as a blueprint for similar tools, enhancing preparedness for future public health crises.

## 1. Introduction

The SARS-CoV-2/COVID-19 pandemic originating in Wuhan, China, started in late 2019 and quickly escalated into a global health crisis. Nations worldwide responded with stringent mitigation measures to limit virus transmission, alleviate the healthcare burden, and reduce COVID-19-related fatalities. Simultaneously, substantial efforts were dedicated to prioritising the development and widespread administration of COVID-19 vaccines as soon as they were approved and became readily available. Soon after their development (in late 2020 to early 2021), the European Medicines Agency (EMA) granted conditional marketing authorisation for five vaccines to prevent severe COVID-19 disease and to lower transmission in the European Union (EU) [1]. These approved vaccines included the Comirnaty (previously known as Pfizer-BioNTech) and Spikevax (Moderna) mRNA vaccines, the Vaxzevria (Oxford-AstraZeneca) and Jcovden (Johnson & Johnson) viral vector vaccines, and the protein-based Nuvaxovid (Novavax) vaccine [2].

Large-scale clinical trials confirmed the effectiveness of these vaccines [3]. However, following their necessary rapid conditional introduction, these vaccines were subject to pharmacovigilance monitoring to identify potential rare but serious side effects [4]. In early 2021, a safety signal on the risk of Thrombotic Thrombocytopenia Syndrome (TTS), associated with the viral vector vaccine Vaxzevria was identified, followed by signals of risk of myocarditis (inflammation of the heart muscle) or pericarditis (inflammation of the lining surrounding the heart) associated with the mRNA vaccines (Comirnaty and Spikevax vaccines). For both signals, a careful assessment of the available evidence at the time led to warnings and an update of the product information of these vaccines [4,5]. In response to concerns about the potential risks associated with the COVID-19 vaccines [6], the EMA highlighted opportunities to improve several aspects of vaccine benefit-risk assessment, including data availability and methodologies to enhance contextualisation. Accordingly, the BRAVE (Benefit Risk Assessment of VaccinEs) toolkit was developed, to enable flexible and comprehensive assessments, thereby enhancing the understanding of the risks and benefits associated with COVID-19 vaccination in the European Union. The tool utilises real-time, virus-related clinical data (confirmed infections, hospitalisations, Intensive Care Unit (ICU) admissions, and deaths) alongside estimates of vaccine effectiveness and vaccine risk incidence rates to contrast estimated benefits and risks of vaccination. The results from the analysis using this tool confirmed the overall positive benefit risk balance of Vaxzevria regarding the risk of TTS compared to its benefits, which include confirmed reductions in infections, hospitalisations, ICU admissions, and deaths [5].

Subsequently, suspected signals of myocarditis or pericarditis risks emerged with the extensive deployment of the mRNA vaccines for COVID-19, prompting further updates and warnings [7]. These rare conditions have triggered scientific and societal debates regarding the safety of mRNA vaccines, particularly given that this marks the first large-scale utilisation of the mRNA vaccine platform. Therefore, there is a pressing need for enhanced safety monitoring to address the potential occurrence of these adverse events. Given the increased risk of myocarditis and pericarditis associated with COVID-19, it is essential to compare the incidence rates of these conditions in both vaccinated individuals and those infected with COVID-19 [6]. However, unlike vaccine-associated TTS, for which no background incidence rate in an unvaccinated population was available, there is data on the background incidence of myocarditis/pericarditis. The risk assessment thus involves comparing the observed incidence rates against a reliable estimate of the expected incidence rate. This comprehensive approach ensures a balanced understanding of the benefits and risks associated with mRNA vaccination, ultimately guiding public health decisions and policy-making.

In this study, we aim to demonstrate the use of the BRAVE toolkit for the benefit-risk contextualization of the mRNA vaccines, Comirnaty and Spikevax, by enhancing the quantification and visualisation of vaccine benefits, thereby facilitating their comparison with associated risks. We also demonstrated how easily the vaccine effectiveness parameters in the toolkit could be updated from those employed by Dorta et al [5]. We conducted a comprehensive assessment of age-specific profiles of vaccine benefits (confirmed infections, hospitalizations, ICU admissions, and deaths) and risks of myocarditis or pericarditis within 14 days after vaccine administration for these mRNA COVID-19 vaccines—Comirnaty, authorised across the European Union on December 21, 2020, and Spikevax, authorised on January 6, 2021. This assessment was carried out across various age groups between December 13, 2020, and December 31, 2021. Moreover, we expand the output of the BRAVE toolkit by including uncertainty assessments of the estimated benefits associated with vaccination. This augmentation enables us to account for uncertainties related to the estimation of vaccine effectiveness and further strengthens the toolkit’s utility in informing decisions during future public health emergencies.

## 2. Methodology

The BRAVE toolkit quantifies the benefits and risks of vaccination and requires information on COVID-19 clinical events (confirmed infections, hospitalisations, ICU admissions, or deaths), SARS-CoV-2 variants of concern (VoCs) and vaccine effectiveness estimates for the benefits and information on observed risk events post-vaccination and background risk incidence rates for the risk quantification. The clinical and risk events can be categorised by demographics (i.e., age, sex, geographic location). For the benefit-risk assessment of the mRNA vaccines, the risks are categorised by age and sex, whereas benefits are categorised by age only, as no sex-specific estimates of vaccine effectiveness are available. A brief explanation of the minimal required data for the benefit-risk assessment with the toolkit is summarised in Table S1 of the Supplementary Material.

### 2.1. An extension of the probabilistic model quantifying benefit

The implementation of the probabilistic model in the BRAVE toolkit is based on the following logic. Let *n*(*a,t)* denote the frequency of a specific clinical event *B* (i.e., either confirmed SAR-CoV-2 infections, hospitalisations, ICU admissions, or deaths) at time *t* and for a specific age group *a*. The calculation of the (counterfactual) frequency of clinical event *B* without vaccination is as follows:

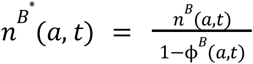

where *ϕ*_*B*_(*a,t)* denotes the proportion of individuals in age group *a* protected through vaccination at time *t* for clinical event *B*. In this analysis, we assume no delay between SARS-CoV-2 infection and the occurrence of the clinical event. This simplification, though unrealistic, has a negligible impact on results due to the slow buildup of protection induced by vaccination compared to the maximum delay between infection and for example confirmation of infection [5].

The proportion of vaccinated individuals in age group *a* at time *t* with vaccine dose *l* can be defined as an accumulation of the proportion of vaccinated individuals 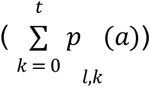 at a discrete time point (with time steps of one day) *k* = 0, 1, 2, …,*t*, with *k* = 0 representing the first day of vaccine administration. A correction factor (denoted by 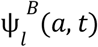) for vaccine dose *l* = 1, 2 was considered to accommodate changes in vaccine effectiveness based on the prevalence and a time– and age-invariant correction factor for each variant (original, α and δ), as well as partial protection resulting from the first dose of a COVID-19 vaccine (assuming both vaccines are mRNA-based) when assessing the benefits of vaccination. The vaccine effectiveness concerning clinical event *B* in relation to the time since vaccination *t* – *k* is defined by 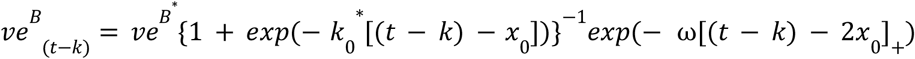, with *x*_0_ representing the midpoint of a logistic growth curve. Here, we assume that vaccine effectiveness exhibits logistic growth after vaccination, reaching a level of protection equal to 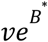, after which an exponential decay takes place with a waning rate equal to ω [5]. The operator [.]+ equals zero if the argument is negative and takes the value of the argument when positive. To calculate the proportion of protected individuals in age group *a* at time *t*, we weigh the contribution of the overall protection, by relying on the proportion of individuals of this cohort being vaccinated only once or twice in the population. Let *w*_1,*t*_ represents the proportion of individuals in age group *a* at time *t* who have only received the first dose of vaccination, hence it can be written as

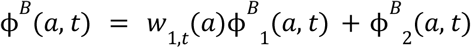

With

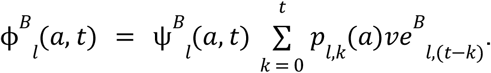

Extending this BRAVE toolkit, we employ Monte Carlo simulations to incorporate uncertainty into the probabilistic model [8], assuming independence of the vaccine benefits across countries. This means that the benefits of a country’s vaccination program are not influenced by those observed in other countries. Here, we conducted 1,000 independent simulations, sampling vaccine effectiveness parameters from uniform distributions across specified intervals. The results were then aggregated to construct percentile-based 95% confidence intervals for key population quantities, such as the mean number of averted COVID-19 related confirmed infections. In this exercise, we consider the variability arising from the vaccine effectiveness of Spikevax and Comirnaty for each COVID-19 burden per dose and variants of concern **(Table S2)**. Note that this extension is conducted outside of the publicly available toolkit due to its computational burden. Detailed information on vaccine uptake and age– and time-specific event occurrence data is crucial for determining vaccine benefits. For countries lacking specific information, we applied a multiple imputation approach within the probabilistic model framework, similar to the method used by Dorta et al [5].

### 2.2. Risk quantification with Observed-Expected (O/E) ratio

The risk ratio per age group *a* and sex *s* is calculated by comparing the joined observed myocarditis and pericarditis events with the background incidence rates,

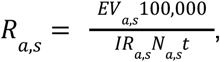

Where *IR*_*a,s*_ represents the background incidence rates for myocarditis and pericarditis per sex and age, expressed per 100,000 person-years; *t* denotes the time horizon in years; and *EV*_*a,s*_ refers to the observed myocarditis and pericarditis events associated with mRNA COVID-19 vaccination within a 14-day (0.093 years) post-vaccination time interval, as reported to the European Union Drug Regulating Authorities Pharmacovigilance **(Table S3)**. A small subset of these data lacked age and/or sex information, which was remedied by imputation with referencing proportions from fully documented EudraVigilance cases. The vaccination coverage per sex and age (*N*_*a,s*_) combined different data sources and required a redistribution of observed coverages to the risk-appropriate age-by-sex categories. As a sensitivity analysis, two types of adjustments were applied, namely redistribution via fixed proportions or redistribution via multiple imputation, with the latter being considered most appropriate.

The background incidence rates for myocarditis and pericarditis jointly were obtained from m = 3 databases (*i* = 1,2,3), one from the *Agenzia Regionale di* Sanità *della Toscana* (ARS), covering both primary and secondary care, and two from the *Base de Datos para la Investigación Farmacoepidemiológica en Atención Primaria* (BIFAP), covering primary care only [5]. In each of these datasets, three cohorts of background incidence rates were identified in the target age groups: the pre-pandemic years (2017-2019), the pandemic period before introduction of vaccination, and the pandemic period thereafter. Given that COVID-19 infection may cause myocarditis or pericarditis, we focus on using the pandemic period before the introduction of vaccination as a comparator. The pooled incidence rate from the three data sources and its variance is estimated by weighting the information from these data sources [9]:

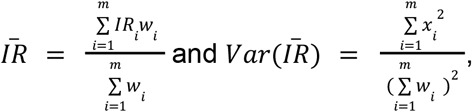

With 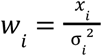 representing the inverse-variance (*σ*_*i*_ ^2^) weighting allowing for a manually chosen contribution of the i-th data source to the mean (*x*_*i*_). The weights should reflect the ability of the data source to estimate the risk incidence rate. Since myocarditis or pericarditis is mainly diagnosed in hospitals and much less in primary care settings, in this study, the ARS data source was given a 10-fold weight compared to each of the primary care BIFAP data sources (both having the same weight). The toolkit thus allows for careful consideration of the fit of purpose and the heterogeneity of the available data sources in estimating risk incidence rates.

## 3. Results

### 3.1. Benefit quantification

Between 13 December 2020 and 31 December 2021, a total of 542.352 million Comirnaty and 108.165 million Spikevax vaccines were administered across 30 EU/EEA countries [10]. We estimated the benefits obtained by administering Comirnaty and Spikevax vaccines **(Table S3)**. In total, the mRNA vaccines prevented an estimated 11.150 million (95% Confidence Interval (CI): 10.876 – 11.345) confirmed COVID-19 infections, 0.739 million (95% CI: 0.727 – 0.743) hospitalisations, 0.107 million (95% CI: 0.104 – 0.108) ICU admissions, and 0.187 million (95% CI: 0.182; 0.189) COVID-19-related deaths across 30 EU/EEA countries during the study period **(Figure 1)**.

**Figure 1:**
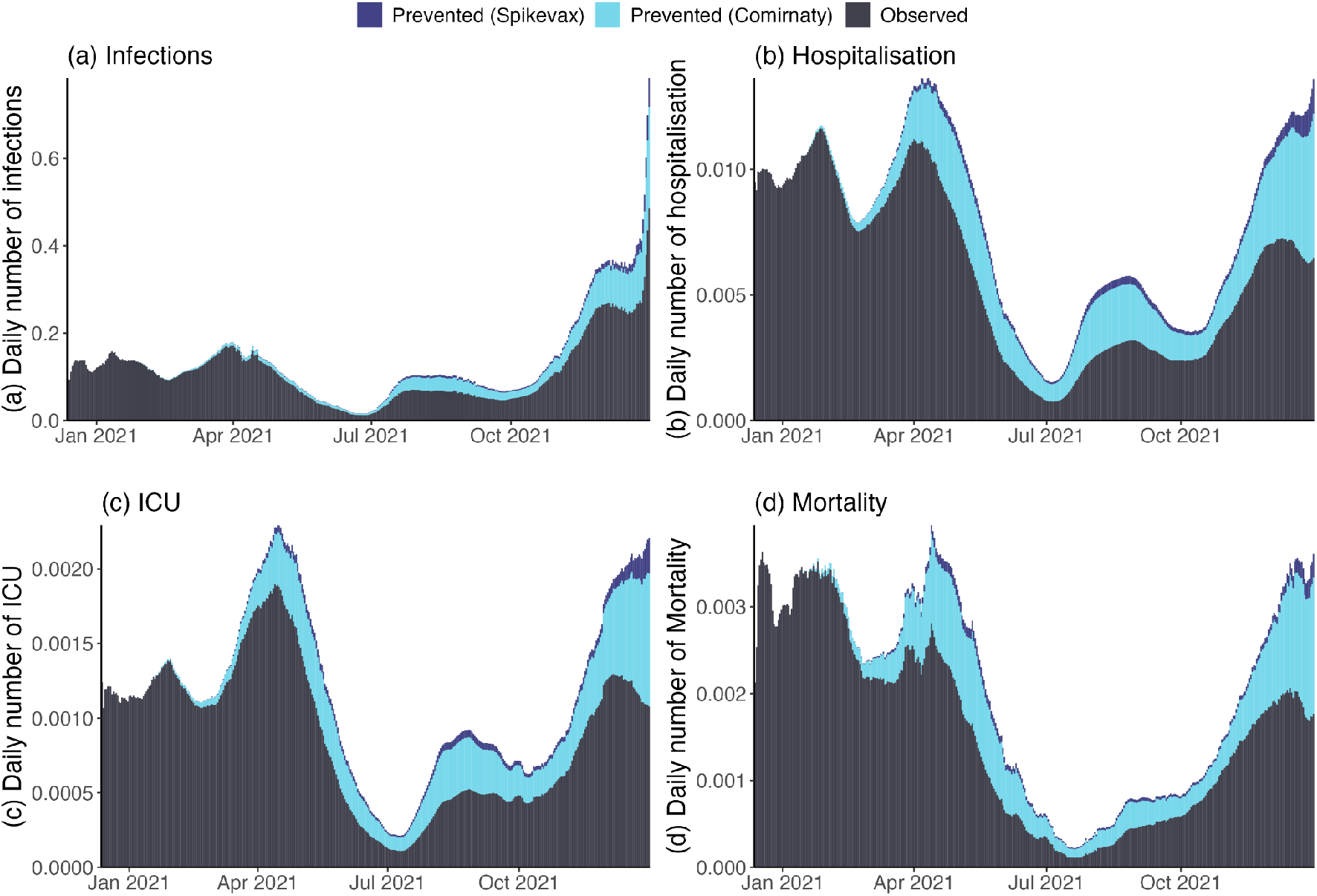
Daily number (in millions) of observed (black) and prevented (light blue: Comirnaty; dark blue: Spikevax) (a) COVID-19 infections, (b) hospitalisations, (c) ICU admissions, and (d) deaths across 30 countries in Europe between December 13, 2020, and December 31, 2021.

During the study period, the mRNA vaccines led to the greatest reduction in COVID-19-related hospitalisations and deaths in the 80+ age group (respectively 347 (95% CI: 339 – 349) per 100,000 administered vaccines and 128 (95% CI: 125 – 130) per 100,000 vaccines), while the 70-79-year-old age group had the greatest number of avoided ICU admissions (41 (95% CI: 40 – 42) per 100,000 vaccines). Additionally, there was a substantial average number of prevented COVID-19 confirmed infections in the 20-59 years age range (1,757 (95% CI: 1,709 – 1,789) per 100,000 vaccines) and the 80+ age group (1,374 (95% CI: 1,351 – 1,392) per 100,000 vaccines), when combining the benefits of Comirnaty and Spikevax vaccines (**Figure 2**). Vaccination was estimated to significantly reduce the number of hospitalisations, ICU admissions, and deaths caused by COVID-19 for age groups above 40 years, with individuals older than 70 years exhibiting the highest number of prevented hospitalisations and deaths due to vaccination **(Table S4)**.

**Figure 2:**
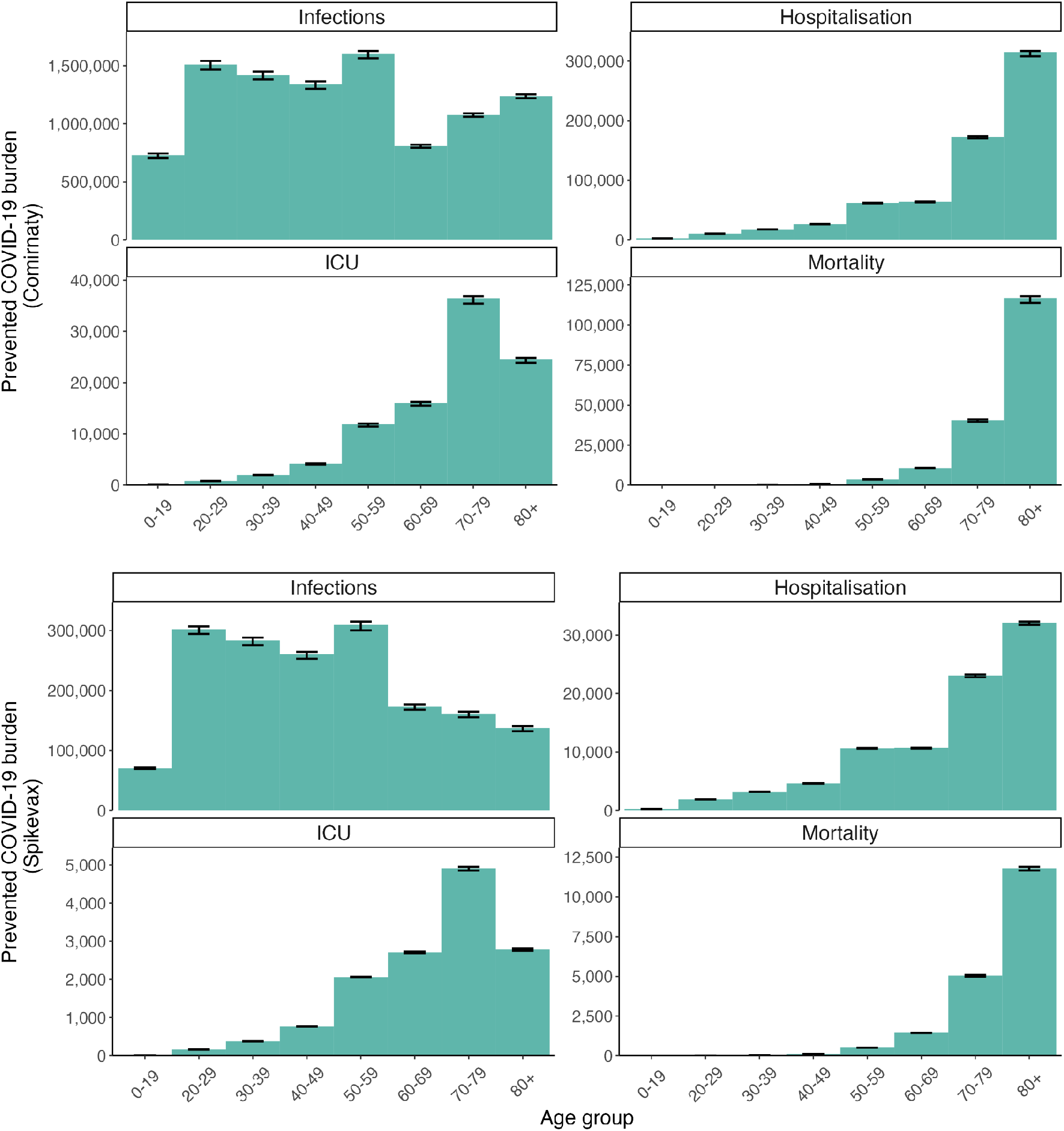
The estimated number (with 95% confidence intervals depicted as error bars in black) of (a) COVID-19 infections, (b) hospitalizations, (c) ICU admissions, and (d) deaths prevented among individuals vaccinated with Comirnaty *(Top)* and Spikevax *(Bottom)* per 100,000 individuals vaccinated in Europe within the respective age categories from December 13, 2020, to December 31, 2021.

### 3.2. Risk quantification

As of October 13, 2021, a total of 4,635 cases of myocarditis or pericarditis potentially related to mRNA vaccination (3,644 cases for Comirnaty and 991 cases for Spikevax) were reported to the European Union Drug Regulating Authorities Pharmacovigilance (EudraVigilance). Across most age categories and for both vaccines, there were more reported myocarditis/pericarditis cases among males than females **(Table S5)**. The Observed-Expected ratios at 14 days post-vaccination estimated using the pooled background incidence rate during COVID-19 though prior to vaccination and relying on multiple imputation, showed higher Observed-Expected ratios in age categories less than 40 years of age for both sexes and vaccine types **(Figure 4)**. Only for Spikevax, the Observed-Expected ratio is above 1, indicating a higher observed number of myocarditis/pericarditis cases than expected in the absence of vaccination. Specifically, an increased risk is observed for males aged between 18 and 24 years, and to a lesser extent, for those aged 25 to 39 years after Spikevax vaccination. In age groups under 40 years, the risk of myocarditis/pericarditis is marginally higher in males than females. The large risk ratio variability observed in females aged 25 to 29 years is attributed to a zero-incidence rate estimate in 2 out of the 3 data sources providing background estimates. A fixed proportion redistribution approach does not change the conclusions, but the multiple imputation redistribution method reduces the variance for the older age categories, while it increases the uncertainty for the younger ones **(Figure S1)**.

**Figure 4:**
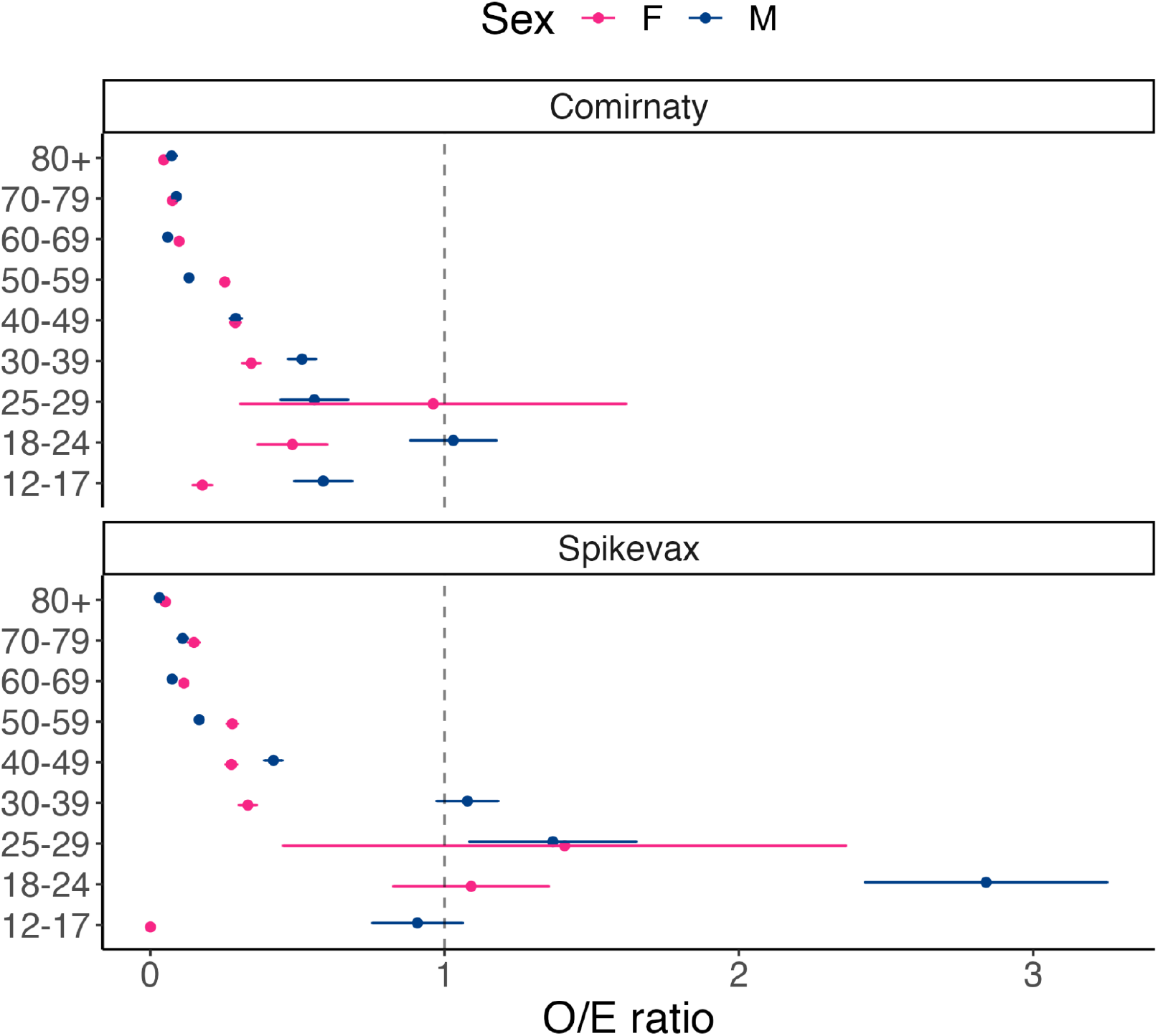
The Observed-Expected ratio of myocarditis/pericarditis cases by age group, sex, and vaccine type (i.e Comirnaty *(Top)*, Spikevax *(Bottom)*), using the pooled background incidence rate during COVID-19 prior to vaccination with weights (1, 0.1, 0.1) and relying on multiple imputation.

## 4. Discussion

We extended the BRAVE toolkit to explicitly accommodate variability with regard to vaccine effectiveness and demonstrated the benefits of COVID-19 mRNA vaccination compared to the risk of myocarditis or pericarditis in 30 EU/EEA countries [5]. The benefits of vaccination (i.e, number of prevented infections, hospitalisations, ICU admissions, and deaths) accounted for varying vaccine effectiveness as a function of time since vaccination, the emergence of new variants of concern and age-specific and temporal differences in disease dynamics and were placed against any vaccination risk, categorised by age and sex subgroups. While acknowledging that the results for Comirnaty and Spikevax vaccines may vary because of differences in timing of vaccination roll-out across countries and in the initial target populations for vaccination, the analysis shows that benefits still far outweigh the potential risks associated with myocarditis and pericarditis within 14 days following vaccination, across all age groups and sexes.

In general, COVID-19 vaccines have shown significant benefits across various age groups [11,12]. Our findings are in alignment with existing research, which consistently shows that COVID-19 vaccination has a beneficial effect in older age groups [13–15]. These age groups are at higher risk of experiencing severe COVID-19-related clinical events and vaccination has played a crucial role in mitigating the impact of the virus in these age groups. Furthermore, younger age groups may also benefit from vaccination by reducing the frequency of infections, thereby reducing the risk of upward transmission to more vulnerable individuals, and protecting themselves from potential long-term effects of the disease [16]. Despite the benefits of COVID-19 vaccination, it is imperative to acknowledge and address the potential risks associated therewith. The findings of the current study align with established evidence from other studies, and earlier benefit-risk assessments by the EMA, highlighting that COVID-19 mRNA vaccine-induced myocarditis or pericarditis is more common among younger males [17,18]. The Observed-Expected ratio for Spikevax consistently exceeded that of Comirnaty, indicating a potentially elevated risk of myocarditis and pericarditis for Spikevax as compared to the Comirnaty vaccine [19–21]. While there was a decrease in the incidence of myocarditis and pericarditis, we observed significantly higher-than-expected values across all age groups. Important to note that our comparison involved the background incidence rate during the COVID-19 period before vaccination, which may have influenced the reporting of myocarditis/pericarditis cases, e.g., changes in healthcare-seeking behaviour or diagnostic practices [22].

The toolkit potentially aids users in quantifying the risks and benefits associated with COVID-19 vaccines. By virtue of its flexibility, the toolkit can be readily augmented and complemented with various functionalities, such as the uncertainty assessment included in this study. Acknowledging uncertainty with respect to input parameters provides a more nuanced perspective on benefits and risks estimated from the data at hand. It also enhances understanding by providing a comprehensive overview of vaccine-related risks and benefits, encompassing a robust methodology, both visually and through direct quantification. Secondly, it facilitates decision-making by presenting the results in a clear and accessible format [23]. Furthermore, this toolkit can serve as a foundation for developing practical digital monitoring applications to monitor the benefits and risks of currently available vaccines in real-life scenarios. By providing population-level insights, this interactive toolkit may ultimately support public health planning for prioritising and optimising interventions and informing regulatory decision-making, making it a valuable asset for assessing COVID-19 vaccination benefits and risks while promoting effective public health communication.

While we have accounted for the uncertainty arising from vaccine effectiveness, we recognise that variability from other sources (e.g., penetration of variants of concern in study populations, extent of waning immunity, incidence rates) can also be substantial [24,25]. However, acknowledging additional uncertainty would greatly increase the computational burden. The ability to capture the full complexity of factors influencing vaccine effectiveness is also limited by data availability. For example, the current probabilistic model only directly quantifies clinical events related to COVID-19 by assuming a constant force of infection across different scenarios. This means that it does not consider indirect effects such as a reduced infection risk as a result of the roll-out of vaccination campaigns. While we have employed best practices and achieved valuable results with the available data, further advancements would require more comprehensive and granular data quality. Additionally, the waning rate is incorporated as the rate at which protection, such as that provided by humoral immunity, diminishes over time. Furthermore, the calculation of benefits does not consider the acquisition of natural immunity in cases where individuals develop immunity through prior infection without vaccination. Consequently, our model may not fully capture the entire spectrum of immune protection scenarios and may underestimate the overall level of immunity within the population [26]. This is also an additional, indirect advantage stemming from COVID-19 vaccination, which can be regarded as an even greater benefit resulting from getting vaccinated against COVID-19. Lastly, risk calculations based on spontaneous reports from EudraVigilance may result in an underestimation of the occurrence of myocarditis or pericarditis in the population under study, and no consideration is given to disease severity.

In conclusion, our analysis extending the BRAVE toolkit further emphasises the substantial benefits of COVID-19 mRNA vaccines relative to their risks. This reinforces the significance of developing user-friendly and flexible toolkits like BRAVE. Such a toolkit enhances the quantification and visualisation of vaccine benefits and associated risks and facilitates extrapolation to guide future decision-making and public health planning initiatives.

## Data Availability

The default datasets can be accessed in the BRAVE toolkit. The full study protocol and report related to this study can be consulted at EUPAS4429. Upon request to the authors, the source code behind the BRAVE toolkit can be provided.

## Contributors

CQ, CC and XK conceived the research questions. All authors participated in the design of the study and analysis plan. JV, JC, NL, LW, GM, NH, and SA conducted different parts of the study. NL, JV, and SA drafted the initial and final versions of the manuscript. All authors critically reviewed early and final versions of the manuscript and results. All authors had access to all data, and HGD, CQ, JV and SA have verified the data. All authors had final responsibility for the decision to submit for publication.

## Data sharing

The default datasets can be accessed in the BRAVE toolkit [5]. The full study protocol and report related to this study can be consulted at EUPAS4429 [27]. Upon request to the authors, the source code behind the BRAVE toolkit can be provided.

## Disclaimer

The views expressed in this article are the personal views of the authors and may not be understood or quoted as being made on behalf of or reflecting the position of the European Medicines Agency or one of its committees or working parties.

