## Supplementary Material for "An extension of the Benefit Risk Assessment of VaccinEs (BRAVE) Toolkit to evaluate Comirnaty and Spikevax vaccination in the European Union"

* Shared first authorship
** Shared last authorship

### 1. Minimal data requirements for the BRAVE Toolkit

The minimum data required to run the BRAVE tool includes COVID-19 case data (number of cases, hospitalisations, ICU admissions, mortality), information on SARS-CoV-2 variants of concern (VoCs), and COVID-19 incidence stratified by relevant demographics (e.g., country, age group, vaccine type), as well as estimates of vaccine effectiveness for different doses (Table S1). For our study, the data utilised covers the period from 13 December 2020 to 31 December 2021 which comes from various sources. Here, we briefly describe the data utilised to perform the analyses with the BRAVE toolkit, with details available in Hector et al [1].

***EU/EEA country demographics***
The demographic dataset, obtained from EUROSTAT's December 2021 report, includes population data for each country, categorised by 10-year age groups.

***Granular COVID-19 incidence***
The study utilised datasets obtained from the European Medicines Agency (EMA) and the European Centre for Disease Prevention and Control (ECDC), encompassing COVID-19 data such as confirmed cases, hospitalisations, ICU admissions, and mortality. These datasets spanned 20 EU/EEA countries and covered the period from January 2020 to March 2021.

***Aggregated COVID-19 incidence***
COVID-19 incidence data for cases, ICU admissions, hospitalisations, and mortality were aggregated daily by country from February 26, 2020, to February 9, 2022.

***Variants of Concern (VoCs)***

The percentage of each Variant of Concern (VoC) among the sequenced SARS-CoV-2 positive samples, categorised by both day and country, obtained from the GISAID EpiCOV genomic surveillance database and the European Surveillance System (TESSy) as of May 11, 2020.

***Detailed vaccination coverage and effectiveness***

The vaccination coverage data were sourced from the ECDC Vaccine Tracker submissions to TESSy dd. 10 February 2021 and the COVID-19 vaccine roll-out from the EU/EEA member states by day, vaccine brand, country, 10-year age categories, and the number of doses received. Estimates of vaccine effectiveness (together with the 95% Confidence Interval) for different doses were obtained from different literature and are outlined in Table S2.

***Observed events***

Post-vaccination risk events stratified by vaccine type, age group, and gender of the vaccinated individuals were derived from spontaneous case reports collected within the European Union Drug Regulating Authorities Pharmacovigilance (EudraVigilance database) (data retrieved July 25th, 2021).

***Background incidence rates***

As of November 10th, 2021, the EMA provided background incidence rates (IR) for myocarditis and pericarditis. This dataset offers insights into the incidence of these conditions across different age groups and genders, detailing the number of cases observed relative to person-years for two distinct periods: the pre-pandemic period and the period before the initiation of vaccination campaigns during the COVID-19 pandemic.

***Aggregated vaccination coverage***

The aggregated vaccination coverage data includes information on coverage rates per country, gender, and age group. In instances where the data lacks gender and/or age details, it undergoes redistribution using two methodologies. The first method involves fixed proportions, while the second incorporates uncertainty around the proportions through multiple imputations. Users can choose between these methodologies within the toolkit interface.

### 2. Additional Figures

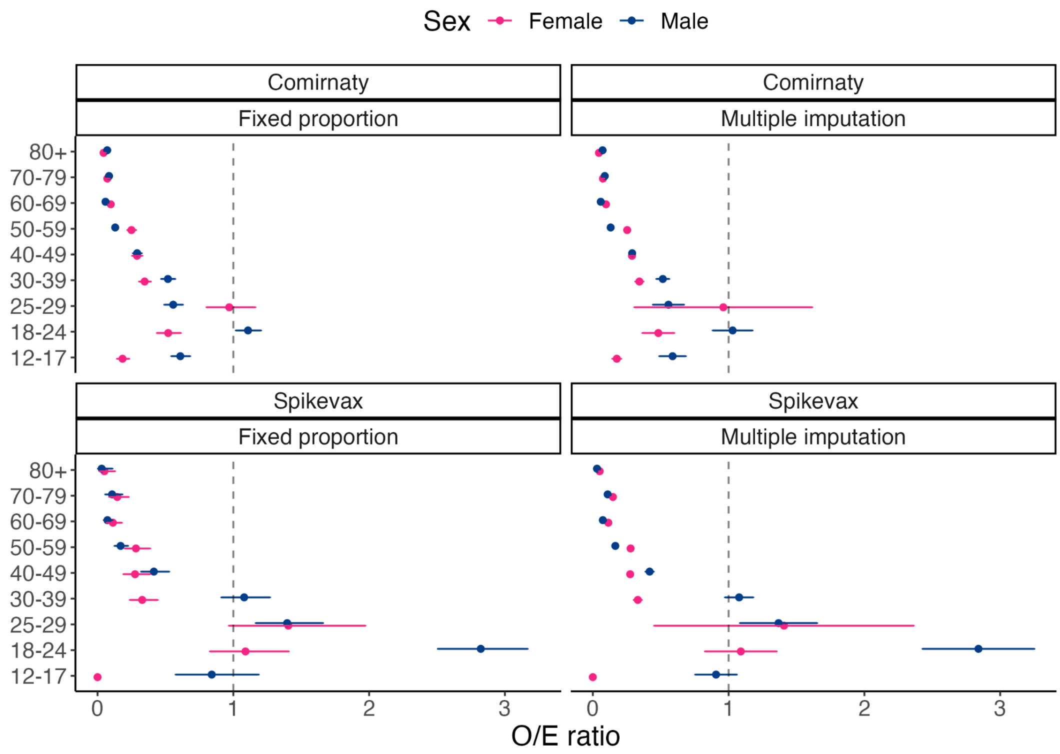


**Figure S1:** The Observed-Expected ratio of myocarditis/pericarditis by age, vaccine type and sex (*Top*: Comirnaty; *Bottom*: Spikevax), using the pooled background incidence rate during COVID-19 prior to vaccination and with weights (1,0.1,0.1) comparing *(Left)* fixed proportion and *(Right)* multiple imputation redistribution of the vaccine coverage.

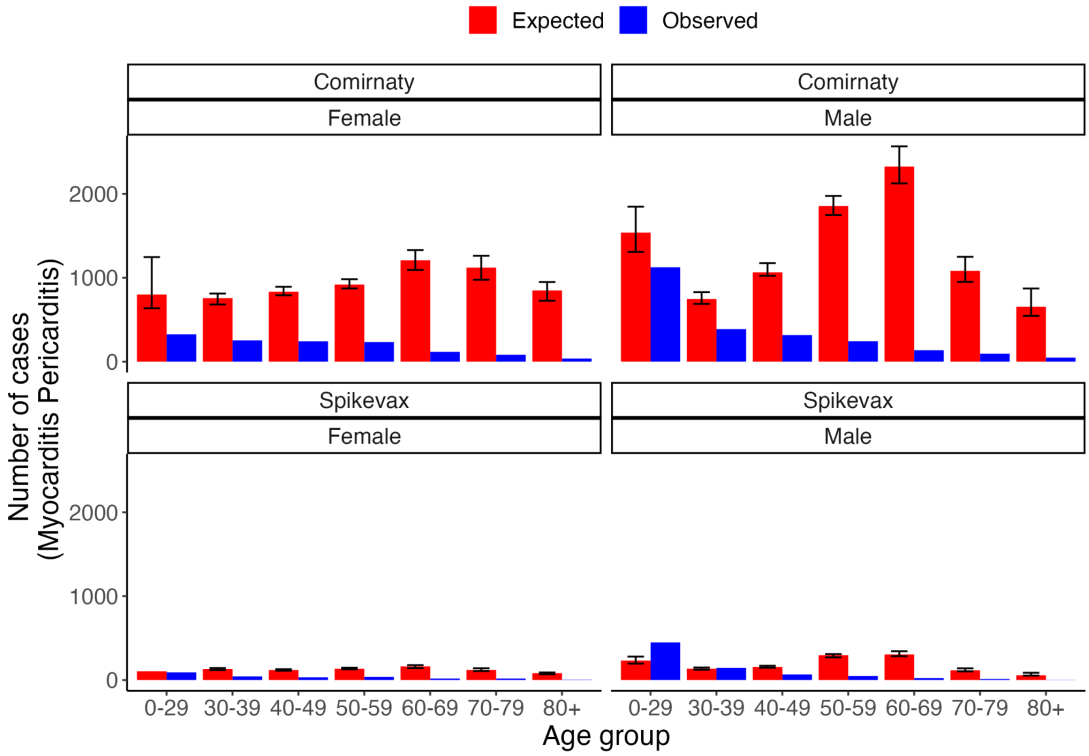


**Figure S2:** The number of expected and observed risks (myocarditis/pericarditis) by age and sex, associated with Comirnaty *(Top)* and Spikevax *(Bottom)* vaccination during the study period.

### 3. Additional Tables

**Table S1:** Required data for the benefit risk contextualisation in the BRAVE toolkit. The COVID-19 clinical endpoint pertains to the count of cases, hospitalisations, ICU admissions, or deaths resulting from the COVID-19 virus.

| No. | Benefits datasets | Risks datasets |
|---|---|---|
| 1 | Population size (by age/gender) for different EU countries | Number of observed events for the risk of interest aggregated by population of interest |
| 2 | COVID-19-related clinical endpoint by population of interest aggregated by date. | Background incidence rates of the reported risk of interest aggregated by the population of interest |
| 3 | COVID-19-related clinical endpoint by the population of interest. | Vaccine coverage aggregated by population of interest |
| 4 | Prevalence of SARS-COV-2 variant of concerns (VOCs) | |
| 5 | Age- and Time-dependent vaccine coverage aggregated by population of interest. | |

**Table S2:** Overview of the default parameters used in the analysis and toolkit. These parameter values correspond to $ve^*$, $ve^*_\alpha$, $ve^*_\delta$ parameters mentioned in the main text (allowing for differential protection depending on the vaccine dose). In cases where detailed information is lacking for specific clinical outcomes, we consider parameter values from less severe endpoints in the following order: infection, hospitalisation, ICU admission, and mortality [8].

| Vaccine type | Vaccine dose | Variant | Clinical outcome | Value (95% CI) | Source |
|---|---|---|---|---|---|
| Comirnaty | 1 | Alpha | Infection | 0.48 (0.42 - 0.53) | Lopez et al (2021)b [2] |
| | | Delta | | 0.36 (0.23 - 0.46) | Lopez et al (2021)b [2] |
| | | Wuhan | | 0.52 (0.30 - 0.69) | Creech et al (2021) [3] |
| | | Alpha | Hospitalisation | 0.86 (0.83 - 0.89) | Andrews et al (2022)b [4] |
| | | Delta | | 0.91 (0.90 - 0.92) | Andrews et al (2022)b [4] |
| | | Wuhan | | 0.89 (0.20 - 0.99) | Creech et al (2021) [3] |
| | | Alpha | ICU admission | 0.86 (0.83 - 0.89) | |
| | | Delta | | 0.91 (0.90 - 0.92) | |
| | | Wuhan | | 0.89 (0.20 - 0.99) | |
| | | Alpha | Death | 0.89 (0.79 - 0.94) | Andrews et al (2022)b [4] |
| | | Delta | | 0.89 (0.79 - 0.94) | Andrews et al (2022)b [4] |
| | | Wuhan | | 0.89 (0.20 - 0.99) | |
| | 2 | Alpha | Infection | 0.94 (0.92 - 0.95) | Lopez et al (2021)b [5] |
| | | Delta | | 0.88 (0.85 - 0.90) | Lopez et al (2021)b [5] |
| | | Wuhan | | 0.95 (0.90 - 0.97) | Creech et al (2021) [3] |
| | | Alpha | Hospitalisation | 0.98 (0.91 - 0.99) | Andrews et al (2022)b [4] |
| | | Delta | | 0.99 (0.98 - 0.99) | Andrews et al (2022)b [4] |
| | | Wuhan | | 0.95 (0.90 - 0.97) | |
| | | Alpha | ICU admission | 0.98 (0.91 - 0.99) | |
| | | Delta | | 0.96 (0.90 - 0.98) | Price et al (2022) [6] |
| | | Wuhan | | 0.95 (0.90 - 0.97) | |
| | | Alpha | | 0.96 (0.94 - 0.97) | Andrews et al (2022)b [4] |

| | | | | | |
|---|---|---|---|---|---|
| | | Delta | Death | 0.99 (0.97 - 0.99) | Andrews et al (2022)b [4] |
| | | Wuhan | | 0.95 (0.90 - 0.97) | |
| Spikevax | 1 | Alpha | Infection | 0.58 (0.12 - 0.80) | Andrews et al (2022)b [4] |
| | | Delta | | 0.65 (0.64 - 0.66) | Andrews et al (2022)b [4] |
| | | Wuhan | | 0.92 (0.69 - 0.99) | Creech et al (2021) [3] |
| | | Alpha | Hospitalisation | 0.94 (0.90 - 0.96) | Andrews et al (2022)b [4] |
| | | Delta | | 0.94 (0.90 - 0.96) | Andrews et al (2022)b [4] |
| | | Wuhan | | 0.92 (0.69 - 0.99) | |
| | | Alpha | ICU admission | 0.94 (0.90 - 0.96) | |
| | | Delta | | 0.94 (0.90 - 0.96) | |
| | | Wuhan | | 0.92 (0.69 - 0.96) | |
| | | Alpha | Death | 0.94 (0.90 - 0.96) | |
| | | Delta | | 0.94 (0.90 - 0.96) | |
| | | Wuhan | | 0.92 (0.69 - 0.99) | |
| | 2 | Alpha | Infection | 0.98 (0.97 - 0.99) | Bruxvoort et al (2021) [7] |
| | | Delta | | 0.87 (0.84 - 0.89) | Bruxvoort et al (2021) [7] |
| | | Wuhan | | 0.94 (0.90 - 0.97) | Creech et al (2021) [3] |
| | | Alpha | Hospitalisation | 0.98 (0.97 - 0.99) | |
| | | Delta | | 1.00 (1.00 - 1.00) | Andrews et al (2022)b [4] |
| | | Wuhan | | 0.98 (0.93 - 0.99) | Bruxvoort et al (2021) |
| | | Alpha | ICU admission | 0.98 (0.97 - 0.99) | |
| | | Delta | | 1.00 (1.00 - 1.00) | |
| | | Wuhan | | 0.98 (0.93 - 0.99) | |
| | | Alpha | Death | 0.98 (0.97 - 0.99) | |
| | | Delta | | 1.00 (1.00 - 1.00) | |
| | | Wuhan | | 0.98 (0.93 - 0.99) | |

**Table S3:** Estimated benefits obtained by the BRAVE toolkit per vaccine type.

| | **Comirnaty** | **Spikevax** |
|---|---|---|
| Confirmed COVID-19 infections | 9.47 million [9.25 - 9.64] | 1.68 million [1.62 - 1.70] |
| Hospitalisations | 0.65 million [9.25 - 9.64] | 0.08 million [0.08 - 0.08] |
| ICU admissions | 0.09 million [9.25 - 9.64] | 0.01 million [0.01 - 0.01] |
| Deaths | 0.17 million [9.25 - 9.64] | 0.02 million [0.02 - 0.02] |

**Table S4:** The number of prevented disease burdens by age, including prevented clinical events, associated with Comirnaty and Spikevax vaccination combined during the study period.

| **Age group** | **Hospitalisations** | **ICU admission** | **Mortality** |
|---|---|---|---|
| 0-29 | 12,428 (12,310 - 12,449) | 970 (934 - 983) | 117 (115 - 118) |
| 30-39 | 20,821 (20,622 - 20,864) | 2,380 (2,302 - 2,405) | 339 (331 - 341) |
| 40-49 | 30,907 (30,607 - 30,974) | 4,939 (4,778 - 4,993) | 933 (913 - 937) |
| 50-59 | 72,633 (71,844 - 72,824) | 13,957 (13,504 - 14,107) | 4,078 (3,987 - 4,102) |
| 60-69 | 74,500 (73,514 - 74,881) | 18,765 (18,224 - 18,974) | 12,172 (11,847 - 12,277) |
| 70-79 | 196,237 (193,120 - 197,393) | 41,420 (40,299 - 41,844) | 45,664 (44,376 - 46,118) |
| 80+ | 347,263 (339,830 - 349,400) | 27,399 (26,612 - 27,651) | 128,751 (125,550 - 129,947) |

**Table S5:** EudraVigilance reported myocarditis or pericarditis cases by gender, age group and vaccine type dd. 13 October 2021. Events are evaluated in a time window of 14 days after vaccination.

| **Myocarditis or Pericarditis** | | | | |
|---|---|---|---|---|
| **Age group** | **Comirnaty** | | **Spikevax** | |
| | Female | Male | Female | Male |
| 12-17 | 69 | 302 | 0 | 32 |
| 18-24 | 140 | 560 | 58 | 288 |

| | | | | |
|---|---|---|---|---|
| 25-29 | 117 | 263 | 33 | 128 |
| 30-39 | 254 | 388 | 43 | 145 |
| 40-49 | 242 | 317 | 33 | 66 |
| 50-59 | 234 | 243 | 38 | 48 |
| 60-69 | 117 | 136 | 18 | 23 |
| 70-79 | 82 | 95 | 18 | 13 |
| 80+ | 37 | 48 | 4 | 2 |
